# Implication of *FOXD2* dysfunction in syndromic congenital anomalies of the kidney and urinary tract (CAKUT)

**DOI:** 10.1101/2023.03.21.23287206

**Authors:** Korbinian M. Riedhammer, Thanh-Minh T. Nguyen, Can Koşukcu, Julia Calzada-Wack, Yong Li, Seha Saygılı, Vera Wimmers, Gwang-Jin Kim, Marialena Chrysanthou, Zeineb Bakey, Markus Kraiger, Adrián Sanz-Moreno, Oana V Amarie, Birgit Rathkolb, Tanja Klein-Rodewald, Lillian Garrett, Sabine M. Hölter, Claudia Seisenberger, Stefan Haug, Susan Marschall, Wolfgang Wurst, Helmut Fuchs, Valerie Gailus-Durner, Matthias Wuttke, Martin Hrabe de Angelis, Jasmina Ćomić, Özlem Akgün Doğan, Yasemin Özlük, Mehmet Taşdemir, Ayşe Ağbaş, Nur Canpolat, Salim Çalışkan, Ruthild Weber, Carsten Bergmann, Cecile Jeanpierre, Sophie Saunier, Tze Y. Lim, Friedhelm Hildebrandt, Bader Alhaddad, Kaman Wu, Dinu Antony, Julia Matschkal, Christian Schaaf, Lutz Renders, Christoph Schmaderer, Thomas Meitinger, Uwe Heemann, Anna Köttgen, Sebastian Arnold, Fatih Ozaltin, Miriam Schmidts, Julia Hoefele

## Abstract

**Background:** Congenital anomalies of the kidney and urinary tract (CAKUT) are the predominant cause for chronic kidney disease below 30 years of age. Many monogenic forms have been discovered mainly due to comprehensive genetic testing like exome sequencing (ES). However, disease-causing variants in known disease-associated genes still only explain a proportion of cases. Aim of this study was to unravel the underlying molecular mechanism of syndromic CAKUT in two multiplex families with presumed autosomal recessive inheritance.

**Methods and Results:** ES in the index individuals revealed two different rare homozygous variants in *FOXD2,* a transcription factor not previously implicated in CAKUT in humans: a frameshift in family 1 and a missense variant in family 2 with family segregation patterns consistent with autosomal-recessive inheritance. CRISPR/Cas9-derived *Foxd2* knock-out (KO) mice presented with bilateral dilated renal pelvis accompanied by renal papilla atrophy while extrarenal features included mandibular, ophthalmologic, and behavioral anomalies, recapitulating the phenotype of humans with *FOXD2* dysfunction. To study the pathomechanism of *FOXD2*-dysfunction-mediated developmental renal defects, in a complementary approach, we generated CRISPR/Cas9-mediated KO of *Foxd2* in ureteric-bud-induced mouse metanephric mesenchyme cells. Transcriptomic analyses revealed enrichment of numerous differentially expressed genes important in renal/urogenital development, including *Pax2* and *Wnt4* as well as gene expression changes indicating a cell identity shift towards a stromal cell identity. Histology of *Foxd2* KO mouse kidneys confirmed increased fibrosis. Further, GWAS data (genome-wide association studies) suggests that *FOXD2* could play a role for maintenance of podocyte integrity during adulthood.

**Conclusions:** In summary, our data implicate that *FOXD2* dysfunction is a very rare cause of autosomal recessive syndromic CAKUT and suggest disturbances of the PAX2-WNT4 cell signaling axis contribute to this phenotype.

## Introduction

Congenital anomalies of the kidney and urinary tract (CAKUT) are the most important cause of renal replacement therapy (RRT) in children aged 0 – 14 years in Europe (41.3%) and the most frequent cause of chronic kidney disease (CKD) up to the age of 30 years.^1, 2^ CAKUT comprises a broad spectrum of malformations of the kidney and urinary tract ranging from vesicoureteral reflux to renal agenesis leading to end-stage kidney failure (ESKF) requiring dialysis and kidney transplantation.^3^ CAKUT can occur either in isolated or syndromic forms.^4^,

Much progress has been made over the last years concerning disease-associated gene identification by using next-generation sequencing (NGS) approaches. Several monogenic forms of CAKUT have been identified, mostly inherited in an autosomal dominant, but also autosomal recessive fashion. Over 45 genes associated with isolated monogenic CAKUT and over 135 genes associate with syndromic CAKUT are known.^6, 7^ Furthermore, copy number variants (CNV) play an important role in CAKUT.^4^ Nevertheless, only about 10 % of CAKUT cases can be genetically solved and incomplete penetrance and variable expressivity are often observed. Monogenic CAKUT is more frequent if severe kidney affection occurs (renal agenesis/dysplasia), in syndromic/familial cases and if there is parental consanguinity.^3, 8^ Judging from knockout (KO) mouse models and familial clustering of CAKUT, there is evidence for further monogenic causes of CAKUT.^3^

Amongst the genes implicated in CAKUT by mouse models are two transcription factors of the Forkhead box (FOX) gene family, *Foxd1* and *Foxd2*. Both genes are highly similar in structure and sequence and likely have partially redundant functions.^9^ No human individuals with CAKUT resulting from *FOXD1* or *FOXD2* disease-causing variants have been described to date. Here, we report the identification of a homozygous frameshift variant and a homozygous missense variant in *FOXD2* in two unrelated families implicated in autosomal recessive syndromic CAKUT with renal hypoplasia, facial dysmorphies, and proteinuria. Systematic phenotyping of *Foxd2* KO mice did not only confirm the renal anomalies previously reported in the literature but additionally recapitulated extrarenal features observed in the affected individuals.^9^ Transcriptome analyses in a *Foxd2* mutant mouse metanephric mesenchyme cell line suggests *Pax2* and *Wnt4* as *Foxd2* downstream targets, providing a putative pathomechanism potentially involving diversion of lineage identity towards renal stroma cells.

## Methods

Detailed methods can be found in the Supplementary Material.

### Human genetics

In case of family 1 (Figure 1A), the study was approved by the local Ethics Committee of the Technical University of Munich (521/16 S) and performed according to the standard of the Helsinki Declaration of 2013. Written informed consent was obtained from all participants or their legal guardians. The family described in this study is part of a larger hereditary kidney disease cohort (“NephroGen”; over 1,000 families), including 313 CAKUT families, located at the Institute of Human Genetics, Technical University of Munich, Munich, Germany.

**Figure 1.**

(A) Pedigree of family 1 and (B) family 2. Solid symbols, affected individuals; circles, females; squares, males; arrows, index and also affected cousin. Double horizontal bars indicate consanguinity. (**C**) **Renal ultrasound of the index individual in family 1 (right kidney). (D-G) Facial photographs of the siblings of family 2 (down-slanting palpebral fissures, deeply set eyes, laterally extended eyebrows, micro-retrognathia, mild ptosis on left eye). (H) Renal biopsy of individual II-2 of family 2.** Note glomerular hypertrophy, mesangial increase and segmental sclerosis (arrows; hematoxylin-eosin stain). **(I) Illustration of FOXD2 protein showing location of both variants. (J) Multiple alignment analysis using CLUSTAL X software (**http://www.clustal.org/clustal2**) demonstrating methionine (M) in position 210 of *FOXD2* is evolutionary conserved. (K) The 3D structural analyses showing the non-covalent interactions between mutated site and its surrounding residues for the p.Met210Val variant in *FOXD2*.** Pink dashes represent clashes between atom pairs, hydrogen bonds are shown in red, polar interactions are colored in orange, green and sky blue dashes represent hydrophobic interactions and van der Waals forces, respectively. The mutant residue Val210 disrupts interactions initially made by the wild-type residue Met210 with the residues Glu206 and Phe223. Chemically stronger bonds are represented for overlapping interactions (i.e. van der Waals forces are not visible due to overlapping with other interactions).

DNA sample of the index case (VI-3) was analyzed by exome sequencing (ES) using a Sure Select Human All Exon 60 Mb V6 Kit (Agilent, Santa Clara, CA, USA) and a HiSeq4000 (Illumina, San Diego, CA, USA) sequencer as previously described.^10^ Other family members were analyzed by targeted Sanger sequencing using an ABI capillary sequencer 3730 (Applied Biosystems, Foster City, CA, USA). In order to identify additional individuals affected by CAKUT and carrying biallelic damaging *FOXD2* variants, we contacted a number of collaborating laboratories (total of 5,000 cases analyzed) and searched GeneMatcher.^11^

In case of family 2 (Figure 1B), identified through GeneMatcher ^11^, all individuals participating in this study were enrolled after obtaining signed informed consent in accordance with human subject research protocols approved by the Ethical Committee of Istanbul University-Cerrahpasa, Cerrahpasa Medical Faculty (No: 139896, Date: 22/10/2020). Microarray studies were performed in affected individuals (II-1 and II-2) using the Illumina Infinium HumanCytoSNP-12 v2.1(300K) microarray chip and Illumina BlueFuse (v.4.5) software. ES was performed in the two affected girls, one unaffected boy, and their parents (i.e., II-1, II-2, II-3, I-1 and I-2, Figure 1B) using an IDT xGen Exome Plus Panel V1.0 kit (Integrated

DNA Technologies, Coralville, IA, USA) and an MGI-T7 (MGI Tech, Shenzhen, China) sequencer. Homozygosity mapping was performed using ES data via HomSI software (Homozygous Stretch Identifier from next-generation sequencing data), which was developed by the Advanced Genomics and Bioinformatics Research Center in the Scientific and Technological Research Council of Türkiye.^12^ Sanger sequencing was performed in all individuals of the family to validate the sequence variation prioritized by ES using an ABI 3130 genetic analyzer (Applied Biosystems, Foster City, CA, USA).

The GeneBank (National Center for Biotechnology Information, NCBI, Bethesda, MD, USA) sequence *FOXD2* NM_004474.4 (corresponding to the canonical Ensembl transcript ENST00000334793.6) was used as reference. NP_004465.3 protein sequence was also selected for further bioinformatic analyses.

### In silico prediction and multiple sequence alignment for the candidate variants, and protein modeling for FOXD2 p.Met210Val (family 2)

*In silico* predictions were performed using Sorting Intolerant From Tolerant (SIFT, https://sift.bii.a-star.edu.sg/), PROVEAN (http://provean.jcvi.org/index.php/), Polymorphism Phenotyping v2 (PolyPhen2, http://genetics.bwh.harvard.edu/pph2/), Mutation Taster (https://www.mutationtaster.org/), Combined Annotation Dependent Depletion (CADD, ttps://cadd.gs.washington.edu/), Rare Exome Variant Ensemble Learner (REVEL),^13^ ClinPred,^14^ and MutPred2.^15^

Multiple sequence protein alignment was performed using Clustal Omega version 1.2.2 (https://www.ebi.ac.uk/Tools/msa/clustalo/). FOXD2 protein structure and PDB files were absent in the RCSB Protein Data Bank (https://www.rcsb.org/). Therefore, the pre-existing model based in AlphaFold database (https://alphafold.ebi.ac.uk/entry/O60548) was used. Gibbs free energy minimization of the wild type protein structure was carried out with RepairPDB command of FoldX software ^16^ with the following syntax: foldx --command=RepairPDB --pdb= AF-O60548-F1-model_v2.pdb --water=CRYSTAL.

Four *in silico* tools, which estimate the impact of missense variants on protein stability, and calculate the changes of unfolding Gibbs free energy, ΔΔG were employed. Protein stability predictions were calculated with DynaMut2 ^17^, INPS-3D ^18^, FoldX ^16^ and PremPS ^19^ using the AlphaFold structure of FOXD2 as input PDB file.

The ΔΔG value between the wild type protein and results of altered amino acid substitution were calculated for the p.Met210Val variant in FOXD2. Finally, 3D structures of the wild type and altered FOXD2 protein structures were generated with DynaMut2. The ΔΔG values (kcal/mol) for the p.Met210Val substitution were interpreted as follows: Predicted ΔΔG<0 is destabilizing in the case of DynaMut2 and INPS-3D. Predicted ΔΔG>0 is destabilizing in the case of FoldX 5.0 and PremPS.

### CRISPR/Cas9 gene editing in mice and mouse phenotyping

Mice were maintained in individually ventilated cages with water and standard mouse chow according to the directive 2010/63/EU, German laws, and German Mouse Clinic (GMC) housing conditions. All tests were approved by the responsible authority of the district government of Upper Bavaria, Germany.

The *Foxd2* KO mouse model was derived using CRISPR/Cas9 technology using the web-based CRISPOR Design Tool^20^ and systematically characterized in the GMC phenotyping screen as described previously.^21, 22^ Homozygous *Foxd2* knockout mice (10 males, 7 females), heterozygous *Foxd2* knockout mice (8 males, 10 females) and wild type controls (33 males, 36 females) were analyzed by the GMC at the Helmholtz Zentrum München, Neuherberg, Germany (http://www.mouseclinic.de).21-23 The phenotypic tests were part of the GMC screen and performed according to standardized protocols as described before.^24–27^ More details and the description of the immunohistochemistry and image analysis can be found in the Supplementary Material.

### Cell culture, Foxd2 deficient metanephric mesenchyme cell models generation using CRSPR/Cas9 and transcriptome analyses

CRISPR/Cas9 gene targeting on mk4 metanephric mouse cells was performed as previously described.^28, 29^ Bioinformatics analysis was performed using the GALAXY platform (https://usegalaxy.org/) as previously described.^30^ qPCR in different mk4 clones carrying different homozygous frameshift alleles to exclude clonal effects was performed using GoTaq qPCR Mastermix (Promega, Madison, WI, USA; Cat. number A6001). Immunofluorescence analyses were done as previously described (Anti-Pax2 antibody: ab79389 [Abcam, Cambridge, UK], 1:200).^31^ Gel electrophoresis and western blotting were likewise performed as previously described (Anti-Pax2 antibody: ab150391, EPR8586 [Abcam]).^32^

### FOXD2 GWAS locus fine-mapping

Genome-wide association studies (GWAS) of albuminuria have identified the *FOXD2* locus.^33^ We therefore attempted to identify the responsible variants underlying the association signal by using the statistical fine-mapping method SuSiE.^34^ SuSiE selects single nucleotide polymorphisms (SNPs) with a high probability to causally affect a given trait - here the urinary albumin-to-creatinine ratio (UACR) - even if a genetic locus contains many highly correlated genetic variants and/or multiple SNPs with a causal effect. We used the R package susieR (version 0.12.27) to fine-map the 1 MB genomic region centered at the SNP showing the strongest statistical association with UACR, rs1337526. For linkage disequilibrium (LD) matrix calculation, we used a genotype set of 15,000 randomly selected participants of European ancestry from the UK Biobank as in Teumer *et al.*.^33^ To match the LD reference as closely as possible, GWAS summary statistics of UACR based on data from 436,392 participants of the UK Biobank (application numbers 20272) was used as input. We used the default parameters in susieR functions, except for setting var_y to 1 and max_iter to 100000. The identified credible set SNPs were positioned in the IGV browser of the human kidney snATAC-seq open chromatin peaks (http://www.susztaklab.com/human_kidney/igv/) from Sheng et al.^35^ to examine their position with respect to cell-type specific open chromatin peaks. The regional association plot was created by LocusZoom version 1.4.^36^

### Code availability

Code for analyses and plottings are available under https://github.com/gwangjinkim/foxd2_analysis.

## Results

### Clinical case

#### Family 1

In this consanguineous family, the index individual (VI-3) had facial dysmorphism (low-set ears, hypertelorism, down-slanting palpebral fissures, retrognathia), developmental delay and congenital bilateral hypoplastic kidneys (Figure 1C) and underwent allogenic kidney transplantation during later childhood. He developed CKD stage 4 (estimated glomerular filtration rate [eGFR] 26 mL/min/1.73m^2^ [modified Schwartz formula]^37^) and proteinuria of 2.7 g/24h at school age. His parents are 1^st^ degree cousins. Three first cousins once removed, individuals V-13, V-14, and V-19 (Figure 1A for a detailed pedigree of the family), were similarly affected. V-19 had bilateral hypoplastic kidneys, developmental delay and retrognathia. At school age, she had CKD stage 3 (eGFR 11 mL/min/1.73m^2^) and proteinuria of 5 g/24h. Allogenic kidney transplantation was performed. Relatives V-13 and V-14, brothers of V-19, were also affected by a similar phenotype (Table 1 and Supplementary Case Report).

**Table 1.**


Overview about the clinical phenotype of the affected individuals of family 1 and family 1. ESKF, end-stage kidney failure; MAF, minor allele frequency; gnomAD, Genome Aggregation Database (v. 2.1.1, https://gnomad.broadinstitute.org); n.d., no data; n.t., not tested.

#### Family 2

In this consanguineous family, the female index individual (II-1) was diagnosed with ESKF at school age. Initial ultrasonography showed bilateral hypoplastic kidneys with increased echogenicity. She received a preemptive kidney transplantation from her father. In addition to kidney failure, the individual had dysmorphic findings including down-slanting palpebral fissures, deeply set eyes, laterally extended eyebrows, micro-retrognathia, and mild ptosis on left eye (Figure 1D-E, Table 1), and high palate, dental crowding, fusiform fingers, sandal gap in both sides, as well as central obesity, noted on physical examination. Eye examination performed at school age showed left esotropia and bilateral posterior subcapsular cataract. The sister of the index individual (II-2) was referred to the pediatric nephrology department due to persistent proteinuria at school age. Her height was 117 cm (−2.61 SD) and she also had dysmorphic findings (Figure 1F-G, Table 1). Serum creatinine level was normal for the age (i.e., 0.35 mg/dL). Renal ultrasound showed bilateral increased renal parenchymal echogenicity and left kidney hypoplasia. Kidney biopsy was compatible with focal segmental glomerulosclerosis (FSGS; Figure 1H). Dysmorphic features included down slanting palpebral fissures, laterally extended eyebrows, micro-retrognathia, left deviation of nasal axis, tapering of distal phalanges of fingers, sandal gap in both sides, short toes, central obesity. In late adolescence, laboratory findings showed impaired renal function (CKD stage 3 with an eGFR of 38 mL/min/1.73 m2 [modified Schwartz formula]^37^).

### Exome sequencing results

#### Family 1

ES of individual VI-3 (index) led to the prioritization of a homozygous frameshift variant in *FOXD2* NM_004474.4:c.789dup, p.(Gly264Argfs*228). The variant is not listed in gnomAD (Genome Aggregation Database, v.2.1.1, http://gnomad.broadinstitute.org/) and predicted to result in a protein of nearly the same length as wild type FOXD2, however, with a changed amino acid composition within the C-terminal half of the protein, leaving the DNA binding domain intact (Figure 1I). As *FOXD2* is a single-exon gene (NM_004474.4), nonsense-mediated decay due to the frameshift variant cannot be expected. See Supplementary Methods for details on filtering process of ES and Supplementary Table 1 for a list of homozygous variants at MAF 1.0% detected with ES in the index individual. Supporting the hypothesis that *FOXD2* represents an essential gene during mammalian development, no *FOXD2* homozygous loss-of-function variants are reported in gnomAD v.2.1.1.

Subsequent Sanger sequencing confirmed segregation of the variant: the parents of VI-3 are heterozygous carriers of the variant. The clinically similarly affected maternal first cousin once removed (V-19) was found to carry the variant homozygously while the maternal great-aunt (IV-6) and mother of V-19 are heterozygous carriers of this variant (Supplementary Figure 1). Further segregation of the variant could not be performed due to limited accessibility/missing consent of relatives.

#### Family 2

Homozygosity mapping revealed runs of homozygosity (ROH) located in chromosome 1, approximately 11.6 Mb long (chr1:g.47,607,000-59,155,000, hg19). The identified homozygous stretch showed that both affected siblings were sharing the ROH, however parents and the unaffected sibling were heterozygous for the region of interest (Supplementary Figure 2A). ES was then performed for the index case (II-1), affected and unaffected siblings (II-2 and II-3) as well as healthy consanguineous parents (I-1 and I-2), respectively. ES analyses resulted in the prioritization of a homozygous missense variant NM_004474.4:c.628A>G, p.(Met210Val) in *FOXD2* (predicted as deleterious with CADD, Revel, ClinPred, and MutPred2), which is located within the disease-segregating homozygous region and is not listed in gnomAD (see Supplementary Table 2 for a list of filtered candidate variants detected with ES in the index individual in family 1).

Sanger sequencing results confirmed the p.Met210Val variant in *FOXD2* is segregating with the disease in the family (Supplementary Figure 2B). The homozygous missense variant is located at the methionine residue at amino acid position 210 within the DNA binding domain of FOXD2 (NP_004465.3), which is completely conserved among different species until the level of Danio rerio (Figure 1I-J).

#### Gibbs Free Energy Calculation and the change of 3D protein structure in *FOXD2* missense variant c.628A>G, p.(Met210Val) (family 2)

The starting free energy ΔG for the FOXD2 protein model was 817.623 kcal/mol, and it was lowered to 719.079 kcal/mol to a more stable state after the RepairPDB command of FoldX was applied. 3D protein models for the wild type FOXD2 and the potential changes on the noncovalent bonds caused by the p.Met210Val variant are displayed in Figure 1K. Seven polar, two van der Waals, and four hydrogen bond interactions were reduced to four polar, one van der Waals, and two hydrogen bond interactions after the Methionine at position 210 turned into Valine. All four hydrophobic interactions were lost, and the six clashes on protein structure remained the same after Met210Val alteration (Figure 1K). The p.Met210Val variant decreased protein stability and predicted as “Destabilizing” with all four different protein stability prediction tools upon point mutations. The ΔΔG scores calculated for the variant are displayed in Supplementary Table 3.

#### Fine-mapping of the FOXD2 albuminuria GWAS locus prioritizes two underlying variants

The *FOXD2* locus (1p33) has been previously associated with albuminuria in GWAS meta-analyses of adult study participants.^33^ In agreement, the affected children carrying rare biallelic *FOXD2* variants show significant proteinuria, with one of them even showing the histologic finding of FSGS on kidney biopsy. Fine-mapping of the *FOXD2* locus to identify driver variants of the UACR association signal revealed two common, independent SNPs upstream of *FOXD2*, rs17453832 and rs1337526. Of these, rs17453832 overlaps accessible chromatin regions in the kidney exclusively in podocytes, the relevant cell type for albuminuria of glomerular origin, as observed in FSGS. Together, these findings suggest that rs17453832 represents a regulatory variant of *FOXD2* in podocytes that leads to an association with albuminuria (Figure 2).

**Figure 2.**

From top to bottom: regional association plot, posterior inclusion probability (PIP) plot, RefSeq gene track and eight snATAC-seq peak tracks at the *FOXD2* locus. In the regional association plot, the association p-values from a genome-wide association study of the urinary-albumin-to-creatinine ratio (UACR) from the UK Biobank were plotted on the -log_10_ scale. The correlation (r^2^) of genetic variants in the region with one of the two lead SNPs in the credible sets identified via fine-mapping (rs1337526 and rs17453832) is indicated as color gradients; a genetic variant is assigned to the lead SNP with which is showed stronger correlation. The r^2^ was based on 1000 Genomes EUR genotype data phase 3 November 2014 release. In the PIP plot, only single nucleotide polymorphisms (SNPs) in the 95% credible sets - i.e., the set of SNPs which contains a variant with effect on UACR with a probability greater or equal to 95% - are shown. The color indicates the credible set membership. The vertical axis indicates the PIP value of the SNPs. The eight ATAC-seq open chromatin tracks were from http://www.susztaklab.com/human_kidney/igv/. Endo: endothelial cells, Podo: podocyte, PT: proximal tubule, LOH: loop of Henle, DCT: distal convoluted tubule, CDPC: collecting duct principal cells, CDIC: collecting duct intercalated cells, Immune: immune cells. Two grey vertical dashed lines were placed at positions where credible set SNPs overlap with open chromatin peaks in podocyte, a relevant cell type for albuminuria.

#### Foxd2 deficiency in mice leads to multidevelopmental phenotypes

To investigate the consequences *Foxd2* dysfunction in mice, *Foxd2* KO mice were generated using CRISPR/Cas9 technology and comprehensively phenotyped. RT-PCR and cDNA analysis confirmed absence of *Foxd2* transcript in homozygous mutant animals (Supplementary Figure 3A). Homozygous *Foxd2* KO mice were viable and were born approximately according to Mendelian distribution. Compared to wild type (wt) and heterozygous littermates, more homozygous newborns died shortly after birth (Supplementary Figure 3B).

At 16 weeks, micro-computed tomography (micro-CT) analyses revealed a changed mandible morphology in homozygous *Foxd2* KO mice (Figure 3A-D; n = 4 wt/2 homozygous females and n = 4 wt/4 homozygous males), including an abnormally shortened condylar process and a flattened tip of the coronoid process in male homozygous *Foxd2* KO mice (Figure 3B-D). Importantly, homozygous *Foxd2* KO mice display smaller mandibles (micrognathia or mandibular hypoplasia), as measured with distance 3-5 (Figure 3D).

**Figure 3.**

Mandibular alterations and defective optic disc in *Foxd2* homozygous knock-out (KO) mice. Micro-CT representative images of the skull **(A)** and mandible **(B)** from control (wt) and homozygous (hom) *Foxd2* KO male mice. In **(A)**, the coronoid (left side) and condylar (right side) processes are highlighted by dash line rectangles and in **(B)**, by a magnified inset and arrows, respectively. **(C)** Volume matching of male control (grey) and mutant (red) mandibles at an identical scale allowing easy qualitative comparison of the described morphological changes. **(D)** Mandibular morphometric analysis comparing distance measurements between several anatomical landmarks: 1. dorsal-most point of the coronoid process, 2. antero-dorsal side of the condylar process, 3. ventral-most point of the condylar process, 4. posterior-most point of the angular process, 5. ventral-most point of the front lower part of the mandible. The condylar process is shorter (distances 1-2 and 3-4) and slightly wider (distance 2-3) in mutants than in control animals. These landmarks have already been described in rodents^69^ n = 4 wt/2 hom females and n = 4 wt/4 hom males. Single values and mean values ± SD are shown. Test for statistical significance was not performed due to the low animal number. **(E)** Representative images from eyes of 16 weeks old homozygous *Foxd2* KO mice and age-matched wild type littermates using *en face* OCT modality. *Foxd2*^-/-^ fundus appearance around the optic nerve displays a darker signal, indicating alterations in the optic disc (green arrow). **(F)** SD-OCT images through the optic nerve showed altered optic nerve morphology. 11/17 homozygous *Foxd2* KO mice showed clear optic disc alterations.

Ophthalmic evaluation identified changes in the appearance of the optic disc, imaged by the OCT *en face* modality as a darker ring around the optic nerve head (Figure 3E). SD-OCT images revealed that homozygous *Foxd2* KO mice have posterior deformation of the optic nerve head surface (Figure 3F). We noted different severity in the alterations of the optic nerve in these mice. The total retinal thickness was not significantly altered, only for severely affected eyes a much thinner total retinal thickness was measured (data not shown).

Although macroscopy did not reveal malformations of the urinary system, microscopic analysis by standard hematoxylin-eosin (H&E) staining detected bilateral dilatation of the renal pelvis. Out of six homozygous Foxd2 KO mice examined (4 males, 2 females), bilateral dilatation of the renal pelvis was present in two homozygous males (33% penetrance; Figure 4A). A clearly narrowed cortex and a decreased size of the renal papilla (Figure 4A+B) accompanied the dilatation. For better visualization of the papilla, we used immunohistochemical staining with Aquaporin2 as a marker for the collecting tubules, and detected a reduction in the diameter of the ducts (Figure 4C). To rule out that the bilateral dilations of the renal pelvis observed in homozygous mutants represented a background lesion, we performed H&E staining of kidneys from 126 wild-type (wt) animals randomized by age and background strain. We detected bilateral renal pelvis dilatation in only four of 126 wt animals (Fisher’s exact test, *p* = 0.0239). Taken together, our results support the diagnosis of a renal pelvis dilation in homozygous mutant mice with a penetrance of 33%. The bilateral dilatation of the renal pelvis was apparently not secondary to an obstructive lower urinary tract lesion. Additional histological parameters of the kidneys are described in Supplementary Table 4 A+B and Supplementary Figure 4.

**Figure 4.**

Histopathological renal alterations in 16-week-old mice. **(A)** Representative overview pictures of hematoxylin-eosin (H&E) staining from left and right kidney of wild type (wt) and *Foxd2* homozygous knockout (KO) mice are shown. Note the normal kidney morphology observed in wt animal compared with bilateral mild renal pelvis dilatation (marked using asterisk) in the homozygous KO mouse. Note the reduced size of the renal papilla (marked in red) and the cortex, which is, clearly narrowed (yellow). **(B**) H&E stains show higher magnification of the renal papilla and immunohistochemistry (IHC-)Aquaporin in **(C)** shows clearly reduction in diameter of the collecting ducts in the mutant mouse. **(D-H)** Plasma clinical chemistry analyses of parameters frequently altered in case of renal dysfunction, including plasma levels of electrolytes (D-F), creatinine (G) and urea (H). Significances according to Mann-Whitney test: * *p* < 0.05; *** *p* < 0.001. hom, homozygous; wt, wild type.

Further, we found a significantly increased (on average doubled) CK8 expression level in the renal cortex of *Foxd2* homozygous knockout mice compared to wt, determined by algorithm-based cell counting (two-sample t-test, *p* < 0.001, Figure 5, Supplementary Figure 5), indicating tubular epithelial injury.

**Figure 5.**

*Foxd2* homozygous knockout (KO) increases cortical CK8 expression. Representative examples of tissue image analysis CK8-staining of renal cortex. Kidney overview with marked cortex and inner medulla of a control (wild type; **A**) and a mutant (homozygous KO**; E)** animal. Magnified renal cortex of a control **(B, C)** and mutant **(F, G)** kidney. Magnified inner medulla of a control **(D)** and mutant **(H)** kidney. Brown = CK8 H-DAB immunohistochemistry. See also Supplementary Figure 5.

Plasma clinical chemistry parameters affected by renal dysfunction showed mild to moderate deviations in homozygous *Foxd2* KO mice compared to wt controls analyzed in parallel. While sodium levels and in trend also chloride values were slightly decreased predominantly in female homozygous *Foxd2* KO mice, maybe as a sign of volume overload due to reduced urine output, a significant increase in plasma potassium and urea levels in male homozygous *Foxd2* KO mice was observed (Figure 4D-F and H). The index individual of family 1 also had increased plasma potassium and urea levels owing to reduced kidney function, but plasma sodium levels were normal. Interestingly, although in general no increase of plasma creatinine concentrations in homozygous *Foxd2* KO mice was observed, two homozygous *Foxd2* KO animals showed high values compared to controls (Figure 4G). One of them was studied by histological analyses and the high creatinine and elevated urea levels were in line with the already described histopathological alterations in the kidneys, including increased cytokeratin 8 expression compatible with increased fibrosis (Figure 5, Supplementary Figure 5).

Behavioral alterations consequent to *Foxd2* loss were also observed. In the open field test of spontaneous reactions to a novel environment, 8-wk-old homozygous *Foxd2* KO mice were clearly hypoactive and hypoexploratory (Supplementary Figure 6). This was indexed by decreased total distance travelled (2-way ANOVA genotype effect: F(1,81) = 19.93, *p* < 0.0001) and total rearing activity (2-way ANOVA genotype effect: F(1,81) = 59.36, *p* < 0.0001) respectively, compared to wt controls. Heterozygous *Foxd2* KO mice were also hypoactive and hypoexploratory compared to wt mice albeit with less severity than in homozygous KO mice (Supplementary Table 5).

#### Generation of Foxd2 deficient metanephric mesenchyme cell models

To further investigate *FOXD2* function for proper renal development and to understand how biallelic *FOXD2* dysfunction may cause bilateral renal hypodysplasia/CAKUT in human and mice, *Foxd2* deficient immortalized mouse metanephric cell models (mk4 cells) were generated using CRISPR/Cas9 technology. mk4 cells represent ureteric-bud-induced metanephric mesenchyme cells undergoing epithelial conversion.

The targeted *FOXD2* region was chosen to closely replicate the homozygous frameshift variant identified in family 1. Four suitable mk4 clones carrying different homozygous *Foxd2* variants resulting in a change of the reading frame were chosen (Table 2, Supplementary Figure 7 [only clone F7 shown]). Foxd2 is a transcription factor previously implicated in renal development in mice ^9^, however, at the time of *Foxd2* KO mouse analysis, RNA seq analysis was unavailable and the precise role for mammalian renal development has remained elusive. Therefore, transcriptome analysis comparing homozygous mutant mk4 to control mk4 cells transfected with non-targeting sgRNA was performed. Clone F7 carrying the frameshift variant NM_00859.3:c.801insT (p.Tyr268Leufs*109) was chosen as the frameshift most closely resembles the one predicted for the allele of the index individual. Five biological replicates per condition were used for transcriptome analysis.

**Table 2.**

*Foxd2* variants created for *in vitro* experiments using CRISPR/Cas9 technology.

#### RNA sequencing results

Transcriptomics analyses revealed a number of differentially regulated genes (Figure 6A). GO term analysis of differentially expressed genes comparing clone F7 to a control clone revealed extracellular matrix organization (GO:0043062, *p* = 3.46=10^-08^)/extracellular matrix (GO:0030198, *p* = 3.378=10^-07^) as top hits followed by several terms related to renal/urogenital development as top hits amongst genes downregulated in the mutant clone: kidney development (GO:0001822, *p* = 5.73=10^-07^), renal system development (GO:0072001, *p* = 6.57=10^-07^) and urogenital development (GO:0001655, *p* = 7.45=10^-07^) with 24-27 out of 6331 genes (adj. *p* = 0.000655 for all three terms). Genes comprised were *Mmp17, Smad9, Fgf1, Emx2, Col4a4, Tfap2a, Sim1, Wnt2b, Adamts1, Tgfb2, Aqp1, Cys1, Pax2, Npnt, Egr1, Agt, Lrp4, Prlr, Col4a3, Igf1, Wnt4, Fgfr2, Id3, Fras1, Gli2, Pygo1*, and *Enpep* (Supplementary Figure 8A-B). Interestingly, amongst upregulated genes in *Foxd2* mutant versus control cells, top GO term hits were related to leucocyte migration, antigen presentation and chemotaxis followed by regulation of MAPK activity (GO:0043405, *p* = 2.96=10^-10^; Table 3, Supplementary Figure 8A-B). Transcriptome analysis of *Foxd2* mutant clone F7 compared to unedited control cells further suggested a significant upregulation of *Nfia* (log2Fc 7,27, adj *p* = 3.358=10^-7^; Figure 6A and Supplementary Figure 9A).

**Figure 6.**

Differential gene expression in *Foxd2* mutant cells. **(A)** Glimma plot visualizing hits of interest. **(B)** Western blot analysis showing strong reduction of Pax2 protein in mutant (clone E4) versus control cells using GAPDH as loading control (n = 3 biological replicates per condition). (**C)** Quantitative analysis of Pax2 levels combining the three biological replicates per condition and GAPDH as a control protein confirming lower Pax2 protein amount in mutant cells compared to controls (Student’s t-test, **** *p* < 0.0001; n.s., not significant).

**Table 3.**

GO-term analysis mk4 clone F7 versus wild type.

qPCR validation on a set of selected targets showed a highly significant correlation with the relative expression levels determined by RNAseq (Pearson R = 0.8955, *p* < 0.0001, Supplementary Figure 9A). In order to exclude clone-specific effects, top gene hits from the GO analysis with potential influence on renal development were confirmed by qPCR, comparing control cells to different *Foxd2* mutant mk4 clones (E4, F6, F7 and H9). These included single gene hits related to early kidney development and/or known to cause renal hypoplasia like *Pax2, Wnt4* and *Fgfr2*. In addition, *Emx2* was found to be downregulated in *Foxd2* mutant mk4 cells compared to control cells. While qPCR confirmed downregulation in all clones compared to control, the log 2 relative expression value (Log2RQ) for *Emx2* was < 1, suggesting a rather minor effect (data not shown). In addition, higher gene expression of *Fat4* in mutant clones was also confirmed compared to wild type. qPCR also confirmed strong downregulation of *Fat2* in all mutant clones compared to control cells (see Supplementary Figure 8C and Supplementary Figure 9B for a full set of qPCR-validated genes).

To investigate *Foxd2* KO on protein level, a Western blot was performed to confirm *Pax2* downregulation. This revealed markedly reduced Pax2 protein levels in Foxd2 deficient cells (three independent experiments with three replicates each per condition, Figure 6B+C).

## Discussion

*FOXD2* encodes a transcription factor of the large, evolutionarily conserved, forkhead gene family important for a variety of processes in humans like organogenesis or metabolism.^38^ *FOX* genes (*FOX* for “forkhead box”) share a “winged helix” DNA binding domain consisting of three N-terminal a-helices, three b-strands and two loops towards the C-terminal region. Different *FOX* genes have been associated with inherited human diseases and cancerogenesis.^39^ Notably, *FOXC1*, a gene associated with syndromic ophthalmologic disease (Axenfeld-Rieger syndrome, type 3; MIM 602482), has recently been linked to autosomal dominant CAKUT.^40^ Additionally, several other FOX genes (*FOXL2*, *FOXA2*, *FOXA3*) have been proposed as candidate genes for monogenic CAKUT in a recent ES study.^41^

However, no representative of the “D” subfamily has been linked to a monogenic disease in human thus far. *FOXD2* (*FREAC-9*, *MF-2*) is well expressed in the kidney cortex.^42^ Of note, the *FOXD2* locus (1p33) has recently been associated with urinary albumin in genome-wide meta-analyses^33^ and *Foxd2* RNA is highly enriched in podocytes and was implicated in maintenance of podocyte integrity.^43^ Proteinuria up to the nephrotic range was reported in the index individual and his female first cousin once removed of family 1 and albuminuria with FSGS on kidney biopsy was present in individual II-2 of family 2 (see Clinical case in the Results section and Supplementary Case Report). In line with this, fine mapping of the *FOXD2* albuminuria GWAS locus revealed a likely regulatory SNP (Figure 2). Together, this could support a role of *FOXD2* for podocyte maintenance and hence in proteinuric kidney disease in general.

Our study suggests that abrogation of *FOXD2* function can result in CAKUT. *Foxd2* KO mice have previously been reported to show renal hypoplasia and hydroureters, however, at reduced penetrance of 40%.^9^ Also, in the KO mice generated for this study, renal anomalies could only be identified at a reduced penetrance of 33% and were rather subtle (Figure 4A-C), underlining the variable expressivity of CAKUT. *Foxd2* shares close sequence homology with *Foxd1* (*Bf2*). A (partial) redundancy of the two could explain the reduced penetrance of CAKUT in *Foxd2* KO mice.^9, 44^ Of note, in contrast to *Foxd2* mutant mice, homozygous *Foxd1* KO mice die shortly after birth of kidney failure due to hypoplastic kidneys.^45^

In Northern blot experiments, mRNA transcripts of *Foxd2* were detected in kidney, facial regions (tongue, nose, maxilla) and brain.^44^ Developmental delay presenting as delayed motor milestone achievement and delayed speech development was noted in the affected individuals of family 1 and 2 (see Clinical case and Supplementary Case Report). Unfortunately, no cranial MRI data were available except for individual II-1 where enlarged ventricles and enlarged subarachnoid space were noted (see Supplementary Case Report). Furthermore, facial anomalies in the affected individuals were compatible with the previously detected expression patterns. A neuronal phenotype was not reported in *Foxd2* KO mice in the literature. However, *Foxd1* KO mice present with small cerebral hemispheres with reduced development of the ventral telencephalon.^9, 46, 47^ Facial alterations, in turn, were not reported in *Foxd2* KO mice in the literature but could be detected in the meticulously phenotyped *Foxd2* KO mice of this study (Figure 3A-D), in line with the facial dysmorphies of the affected individuals of families 1 and 2 (Figure 1D-G; Table 1).^9, 44, 48, 49^ Furthermore, behavioral changes were identified in *Foxd2* KO mice of this study (hypoactivity in the open field test; Supplementary Figure 6). This can be indicative of neurodevelopmental alterations in *Foxd2* KO mice. Taken together, it can be assumed that *FOXD2* plays an important role in neuronal, branchial arch and facial development.

Transcriptome analysis of mk4 CRISPR/Cas9-mediated homozygous *Foxd2* frameshift mutants gave valuable insight into possible mechanisms of CAKUT in the described families. Pathway analysis showed significant enrichment of differentially expressed genes important for development of the renal/urogenital system (Table 3). *Pax2* was downregulated in all mk4 clones (Figure 6A and Supplementary Figure 8C) as were Pax2 protein levels on Western blot (Figure 6B+C). *PAX2* haploinsufficiency is known to be associated with papillorenal syndrome in humans (MIM 120330), which comprises a CAKUT phenotype with eye anomalies.^50^ Of note, no overt eye anomalies were reported in the affected individuals described in this study. However, there is *Foxd2* RNA expression around the eye vesicle in mice during embryonic development.^51^ In line with this, the generated *Foxd2* KO mice showed alterations of optic disc and nerve (Figure 3E+F), which might have been missed on routine ophthalmologic examination of the affected individuals. Interestingly, there were alterations on eye examination of individual II-1 of family 2. However, these did not involve the optic nerve and disc (see Clinical case in the Results section).

*Foxd2* is strongly expressed in renal condensed mesenchyme at embryonic day 11.5 similarly to *Pax2*.^9^ *Pax2* is activated in the mesenchyme in response to induction by the ureteric bud and is subsequently downregulated in more differentiated cells derived from the mesenchyme. With reduced Pax2 protein levels, kidney mesenchyme cells fail to aggregate and do not undergo the sequential morphological changes characteristic of epithelial cell formation, demonstrating an essential role for *Pax2* function for early mesenchymal-epithelial transition (MET).^52^ Interestingly, we also detected a downregulation of *Emx2* in *Foxd2* mutant versus control cells by transcriptomics. *Pax2* enhances *Emx2* gene expression and digenic loss of function of *Pax2* and *Emx2* is known to result in CAKUT similar to what is found in *Foxd2* KO mice and to the phenotype observed in the affected individuals of this study.^9, 53^

Intriguingly, *Pax2* is also implicated in establishing the nephron-interstitium boundary during kidney development: nephron progenitor cells lacking Pax2 fail to differentiate into nephron cells, but can switch fates into Foxd1 positive renal interstitium-like cell types, suggesting that Pax2 function maintains nephron progenitor cells by repressing a renal interstitial cell program.^54^ Our findings suggest that lack of *Foxd2* results in reduced *Pax2* levels. It seems therefore possible that *Foxd2* dysfunction will divert lineage identity towards renal stroma cells. In line with this hypothesis, we could detect significantly increased cytokeratin 8 (CK8) expression in the renal cortex of homozygous *Foxd2* KO mice compared to wt (Figure 5, Supplementary Figure 5). Keratins like CK8 are markers of tubular epithelial injury preceding renal fibrotic changes.^55^ Fittingly, individual II-2 of family 2 featured fibrotic changes on kidney biopsy in terms of FSGS (Figure 1H).

This hypothesis is further supported by the upregulation of *Fat4* in *Foxd2* mutants (Supplementary Figure 8C). *Fat4* encodes an atypical cadherin expressed by stromal cells inhibiting nephron progenitor renewal.^56^ In contrast, there was marked downregulation of *Fat2* in *Foxd2* mutant cells (Figure 6A and Supplementary Figure 8C). While *Fat4* has been shown to play a role in kidney tubule elongation and planar cell polarity in renal cells, the role of *Fat2* for renal development and homeostasis is unclear.^57^ The Drosophila orthologue *fat* has been implicated in cell proliferation and morphogenesis in a contact dependent manner.^58^ We also observed a strong upregulation of *Nfia* in *Foxd2* mutant cells (Figure 6A). Haploinsufficiency of *NFIA* has been associated with brain malformations and CAKUT, and upregulation could be a compensatory mechanism in *Foxd2* KO cells.^59, 60^

In contrast, *Fgfr2* was downregulated in mutant *Foxd2* cells vs. controls (Figure 6A). Conditional KO of *Fgfr2* in metanephric mesenchyme cells leads to CAKUT in mice including hypo-/dysplastic kidneys and hydroureters. Interestingly, in these mice devoid of mesenchymal *Fgfr2* expression, there is no Fgfr2 in stromal cells, either, also indicating a disturbance in stromal cells in *Fgfr2* KO mice.^61^

Further, *Pax2* activates *Wnt4* expression in the metanephric mesenchyme during mammalian kidney development^62^, potentially explaining reduced *Wnt4* gene expression in *Foxd2* mutant cells (Figure 6A and Supplementary Figure 8C). Wnt4 protein plays a role in MET and is essential for tubulogenesis in the developing kidney through a non-canonical Wnt-signaling pathway.^63, 64^ Of note, Wnt4-signalling can be substituted by other Wnt proteins like Wnt7b ^65^ which was also downregulated in *Foxd2* mutant mk4 cells (Supplementary Figure 9B). Figure 7 summarizes the network of assumed *Foxd2* function.

**Figure 7.**

Key players in the network of assumed Foxd2 function. While *Foxd1* expression in the developing mouse kidney is largely restricted to the mesenchymal stroma, *Foxd2* is found in the cap mesenchyme where it plays a role for *Pax2* expression.^9, 70^ *Foxd2* dysfunction leads to reduced PAX2 protein levels (see Results and Figure 6B+C). *Eya1* is important for *Pax2* expression in the UB-induced metanephric mesenchyme (embryonic day 11.5 in mouse) and *Eya1* upregulation in *Foxd2* mutant cells may indicate a compensatory mechanism for reduced Pax2 expression via a feedback loop.^66^ *Pax2* activates *Wnt4* expression in the metanephric mesenchyme during mammalian kidney development and *Wnt4* expression is downregulated in *Foxd2* mutant cells.^62^ *Pax2* enhances *Emx2* gene expression and *Emx2* expression is downregulated in *Foxd2* mutant cells.^53^ Nephron progenitor cells lacking Pax2 can change into Foxd1 positive renal interstitium-like cell types, suggesting that Pax2 represses a renal interstitial cell program.^54^ This is supported by the upregulation of *Fat4* in *Foxd2* mutant cells as *Fat4* encodes an atypical cadherin expressed by stromal cells inhibiting nephron progenitor renewal.^56^ *Fgfr2* (and *Fgfr1*) is believed to act downstream of *Eya1* and upstream of *Pax2* in metanephric mesenchyme.^71^ Conditional KO of *Fgfr2* in metanephric mesenchyme cells leads to CAKUT in mice and deficiency of Fgfr2 in stromal cells.^61^ Figure adapted from ^70, 72^. See Discussion for further details.

In addition, we observed higher *Eya1* expression in *Foxd2* mutant cells compared to control cells (Supplementary Figure 8C). *Eya1* is important for *Pax2* expression in the UB-induced MM and *Eya1* upregulation in *Foxd2* mutant cells may indicate a compensatory mechanism for reduced Pax2 expression via a feedback loop.^66^ *Gdnf* expression, which is vital for UB induction, was unchanged in *Foxd2* mutants (Supplementary Figure 9B). This is in line with *Eya1* upregulation in the context of reduced *Pax2* levels, as both genes regulate *Gdnf* expression.^66, 67^ Hence, it does not seem likely that the renal hypoplasia phenotype observed in the index individuals of the presented families is a result of impaired *Gdnf* expression.

This study has several limitations. The affected individuals of family 1 and 2 do not share the same type of variant. Family 1 segregates a homozygous frameshift variant in *FOXD2*, family 2 features a homozygous missense variant. However, as *FOXD2* is a single-exon gene (NM_004474.4), nonsense-mediated decay cannot be assumed and this makes the frameshift variant not a clear-cut loss-of-function variant but probably leads to an altered protein. Interestingly, no homozygous truncating variants are listed in gnomAD, illustrating that there is constraint for this type of variant in *FOXD2*. This supports a causative role of the frameshift variant. Unfortunately, no patient-derived cells could be obtained in affected individuals of family 1 to clarify FOXD2 expression. This limits the transferability of the KO experiments in this study. On the other hand, it cannot be denied that *Foxd2* KO recapitulates the phenotype of the affected individuals of this study (both of family 1 and 2). And, family 2 also segregates a protein-altering variant - identified by a stringent filtering process - and features striking phenotypic overlap in affected individuals in comparison to family 1. In family 2, it could be shown by *in silico* analysis that the missense variant, located in the DNA binding domain, destabilizes the mature protein. Hence, we believe that both variants abrogate proper FOXD2 function which can be related to by KO experiments. Of course, further studies are needed to experimentally clarify causality of the described variants in *FOXD2* (according to ^68^). Finally, a limitation of our cell culture model is that we investigated similar but not identical *FOXD2* variants *in vitro* compared to the variants we identified in human CAKUT individuals.

In conclusion, our findings indicate that the syndromic CAKUT phenotype in the presented families is caused by *FOXD2* dysfunction, putatively causing a shift of nephron progenitor cells undergoing MET towards a stromal cell identity resulting in fibrotic changes in the kidney. The observed human and *Foxd2* KO mouse phenotype highlights an important role of *Foxd2* in kidney and craniofacial development. The observed kidney alterations are in line with the enrichment of differentially expressed genes important in extracellular matrix organization and renal/urogenital development in *Foxd2* mutant metanephric mesenchyme cells.

It is intriguing that *FOXD2* dysfunction could result in a phenotype of both renal malformation and podocyte damage. As the *FOXD2* locus was also previously associated with urinary albumin in genome-wide meta-analyses^33^ and we have now identified a likely regulatory SNP within this locus, FOXD2 could represent an interesting target in common kidney diseases in terms of tackling proteinuria and renal fibrosis. Consequently, our findings are building bridges between rare monogenic and common complex kidney disease.

## Disclosures

### Ethics approval and consent to participate

This project has received approval from the local Ethics Committee of the Technical University of Munich (521/16 S) as well as from the Ethical Committee of Istanbul University-Cerrahpasa, Cerrahpasa Medical Faculty (No: 139896, Date: 22/10/2020). All participants provided informed consent and the research conformed to the principles of the Helsinki Declaration.

### Consent for publication

Participants whose pedigree data is presented in this manuscript gave consent to publish family history in pedigree or summarized format.

## Data Availability

All data produced in the present study are available upon reasonable request to the authors.

## Acknowledgements

We would like to thank the index individuals and their families for participation in the study and Prof. S. Potter for the kind gift of the mk4 cells. We further thank Prof. Simone Sanna-Cherchi, Prof. Nine Knoers, Dr. Kirsten Renkema and Prof. Sophie Saunier for kindly searching their NGS databases for cases with *FOXD2* variants.

## Supplementary Material table of contents

Supplementary Material includes Supplementary Methods, a Supplementary Case Report, Supplementary Figure 1-9, Supplementary Table 1-5.

## Web Resources

1000 Genomes Project human polymorphism database, http://www.1000genomes.org/

AlphaFold database, https://alphafold.ebi.ac.uk/entry/O60548

ANNOVAR, https://annovar.openbioinformatics.org/en/latest/

Burrows–Wheeler aligner, https://bio-bwa.sourceforge.net/

Combined Annotation Dependent Depletion, CADD, ttps://cadd.gs.washington.edu/ ClinPred, https://sites.google.com/site/clinpred/

ClinVar, https://www.ncbi.nlm.nih.gov/clinvar/

Clustal Omega, https://www.ebi.ac.uk/Tools/msa/clustalo/

Database of Genomic Variants, DGV, http://dgv.tcag.ca/dgv/app/home

dbSNP, http://www.ncbi.nlm.nih.gov/SNP/

DECIPHER, https://decipher.sanger.ac.uk/

DynaMut2, https://bio.tools/dynamut

Expression atlas, https://www.ebi.ac.uk/gxa/home

FoldX, https://foldxsuite.crg.eu/

Galaxy platform12, https://usegalaxy.org/

Genome Analysis Tool Kit, https://gatk.broadinstitute.org/hc/en-us

Genome Aggregation Database (gnomAD v.2.1.1), http://gnomad.broadinstitute.org/

<u>Hom</u>ozygous <u>S</u>tretch Identifier, HomSI, https://bio.tools/homsi

Human Gene Mutation Database, HGMD®, http://www.hgmd.cf.ac.uk

Human Kidney snATAC-seq Peaks, http://www.susztaklab.com/human_kidney/igv/

Leiden Open Variation Database, LOVD, https://www.lovd.nl

INPS-3D, https://inpsmd.biocomp.unibo.it/inpsSuite/default/index3D

Mutation Taster, https://www.mutationtaster.org

MutPred2, http://mutpred.mutdb.org/National

Heart, Lung and Blood Institute Exome Sequencing Project, http://evs.gs.washington.edu/EVS/

Online Mendelian Inheritance in Man^®^, http://www.omim.org/

Picard tool, https://broadinstitute.github.io/picard

PolyPhen2, http://genetics.bwh.harvard.edu/pph2/

PremPS, https://bio.tools/premps

PROVEAN, http://provean.jcvi.org/index.php/

Primer3, http://frodo.wi.mit.edu/primer3/input.htm

QuPath, https://QuPath.github.io/

Rare Exome Variant Ensemble Learner, REVEL, https://sites.google.com/site/revelgenomics/?plix1

RCBS Protein Databank, https://www.rcsb.org/

Sorting Intolerant From Tolerant, SIFT, https://sift.bii.a-star.edu.sg

susieR, https://stephenslab.github.io/susieR/

The Human Protein Atlas, https://www.proteinatlas.org/

UniProt, https://www.uniprot.org/

